# Hypokalemia in Patients with COVID-19

**DOI:** 10.1101/2020.06.14.20131169

**Authors:** Alfano Gaetano, Ferrari Annachiara, Fontana Francesco, Rossella Perrone, Giacomo Mori, Ascione Elisabetta, Magistroni Riccardo, Venturi Giulia, Simone Pederzoli, Gianlulca Margiotta, Marilina Romeo, Francesca Piccinini, Franceschi Giacomo, Volpi Sara, Faltoni Matteo, Ciusa Giacomo, Bacca Erica, Tutone Marco, Raimondi Alessandro, Menozzi Marianna, Franceschini Erica, Cuomo Gianluca, Orlando Gabriella, Santoro Antonella, Di Gaetano Margherita, Puzzolante Cinzia, Carli Federica, Bedini Andrea, Milic Jovana, Meschiari Marianna, Mussini Cristina, Cappelli Gianni, Guaraldi Giovanni, Modena Covid-19 Working Group (MoCo19)

## Abstract

Patients with COVID-19 may experience multiple conditions (e.g., fever, hyperventilation, anorexia, gastroenteritis, acid-base disorder) that may cause electrolyte imbalances. Hypokalemia is a concerning electrolyte disorder that may increase the susceptibility to various kinds of arrhythmia. This study aimed to estimate prevalence, risk factors and outcome of hypokalemia in a cohort of non-critically ill patients. A retrospective analysis was conducted on 290 hospitalized patients with confirmed COVID-19 infection at the tertiary teaching hospital of Modena, Italy.

Hypokalemia (<3.5 mEq/L) was detected in 119 patients (41%). The decrease of serum potassium level was of mild entity (3-3.4 mEq/L) and occurred in association with hypocalcemia (P=0.001) and lower level of serum magnesium (P=0.028) compared to normokaliemic patients. Urine K: creatinine ratio, measured in a small subset of patients (n=45; 36.1%), showed an increase of urinary potassium excretion in the majority of the cases (95.5%). Causes of kaliuria were diuretic therapy (53.4%) and corticosteroids (23.3%). In the remaining patients, urinary potassium loss was associated with normal serum magnesium, low sodium excretion (FENa< 1%) and metabolic alkalosis. Risk factors for hypokalemia were female gender (P=0.002; HR 0.41, 95%CI 0.23-0.73) and diuretic therapy (P=0.027; HR 1.94, 95%CI 1.08-3.48). Hypokalemia, adjusted for sex, age and SOFA score, resulted not associated with ICU admission (P=0.131, 95% CI 0.228-1.212) and in-hospital mortality (P=0.474; 95% CI 0,170-1,324) in our cohort of patients.

Hypokalemia is a frequent disorder in COVID-19 patients and urinary potassium loss may be the main cause of hypokalemia. The disorder was mild in the majority of the patients and was unrelated to poor outcomes. Nevertheless, hypokalemic patients required potassium supplements to dampen the risk of arrhythmias.

## Introduction

COVID-19 is an emerging pandemic spreading worldwide from December 2019. On through 14 June, the death of more than 430.000 people and the massive increase in the number of newly infected people are posing a growing threat to global health[1].

A Chinese epidemiological study documented that the majority of reported COVID-19 cases had a mild presentation (81%) but the development of severe respiratory symptoms (14%) such as dyspnea, tachypnea and hypoxia requires hospital admission.[2] The lung is the principal target of the novel coronavirus and atypical pneumonia is the most common clinical presentation [3]. Multiple organs such as the heart, brain, liver and kidneys can be involved during this infection[4]. Systemic release of cytokines is thought to be the cause of organ dysfunction and, therefore, the major determinant of morbidity in these patients[5]. It provokes a severe pro-inflammatory state leading to hypoxemia and sepsis requiring intensification of supportive therapy. Generally, hospitalized patients belong to a vulnerable subset of the population because they are older adults and have several underlying medical conditions[6]. Patients with COVID-19 can experience a long hospital stay and undergo multiple treatments varying from delivering of ventilatory support to the administration of experimental agents for SARS-CoV-2 infection. In this setting, fever, hyperventilation, sweating, medication-related side effects, change of eating habits may favorite concerning electrolyte imbalance.

Maturing experience in the care of COVID-19 patients showed that hypokalemia is a frequent lab abnormality. There is particular concern about this disorder as it may increase the susceptibility to potential fatal arrhythmia in COVID patients[7,8]. Hypokalemia has been described in a few case-series during the SARS-CoV1 outbreak in 2003, but there is a lack of data on the exact mechanisms underlying this disorder[9,10]. Possible causes of hypokalemia in the context of SARS-CoV2 infection may result from activation of the renin-angiotensin-aldosterone system, gastrointestinal losses, anorexia secondary to concurrent illness and tubular damage caused by ischemic or nephrotoxic kidney damage. Regarding this latter hypothesis, tubular damage may be linked to the direct cytotoxic effect of SARS-CoV-2, since the virus has been associate to diffuse tubular damage[11].

In light of these unanswered clinical questions, we evaluated a cohort of hospitalized patients with the aim to describe the entity of hypokalemia, impact on clinical outcome, and to achieve a better understanding of the underlying pathophysiological mechanism leading this condition.

## Methods

### Study design

A single-center, retrospective, observational study was conducted at the University Hospital of Modena. We retrospectively reviewed all the electronic records of 320 non-critically ill patients admitted to our hospital from February 16, 2020 to April 14, 2020, who were diagnosed with SARS-CoV-2 infection, according to WHO interim guidelines[12]. The study was approved by the regional ethical committee of Emilia Romagna.

### Data collection

We collected data on age, sex, laboratory values during admission (hemoglobin concentration, lymphocyte count, platelet count, arterial blood gas analysis, markers of inflammation, serum and urine creatinine, urea, serum and serum and urine electrolyte), comorbidities, sign and symptoms at presentation, vital signs, administered treatment (diuretics, electrolyte supplement), as well as the admission on ICU and living status at the end of the observation period.

### Outcomes measures

The primary objective of our study was to evaluate the prevalence of hypokalemia. Secondary outcomes included measurement of urinary potassium excretion, the proportion of patients requiring supplemental K, and the impact of hypokalemia on ICU admission and in-hospital mortality compared to normokalemia.

### Variables and definition

Hypokalemia was defined as a serum potassium level < 3.5 mEq/l. The normal level of serum potassium ranges from 3.5 to 5.3 mEq/L. The diagnosis of hypokalemia was performed on a single value of serum potassium < 3.5 meq/L at any time during hospitalization. We excluded all serum values of potassium measured on plasma and by blood gas analyzer. Severity was classified as mild when the serum potassium level was 3 to 3.4 mmol/L, moderate when the serum potassium level was 2.5 to 3 mmol/L, and severe when the serum potassium level was less than 2.5 mmol/L

K urinary excretion was measured as urine K^+^-to-creatinine ratio. A value of K^+^ urinary excretion > 1.5 indicated inappropriate renal potassium loss. K urinary excretion was not measured in patients with chronic and acute kidney disease. Na urinary excretion was measured with the fractional excretion of sodium (FeNa); a value < 1% declared a reduced excretion of sodium, whereas it resulted augmented for value ≥1%. Sequential Organ Failure Assessment (*SOFA*) score was calculated at hospital admission.

### Statistical analysis

Baseline characteristics were analyzed using descriptive statistics and reported as proportions and mean (standard deviation [SD]) when appropriate. Categorical variables were analyzed using χ2 test or Fisher’s test as appropriate. Analyses of continuous variables were compared using an unpaired t-test and Mann-Whitney test, as appropriate. Logistic regression analysis was used to identify the predictors of hypokalemia and to test the risk of in-hospital mortality, adjusted for sex, age and SOFA score at ward admission. A P value of less than 0.05 was considered statistically significant. SPSS 23® was used for statistical analysis.

## Results

### Patients

Serum K^+^ level was measured in 290 (1671 samples) patients with confirmed COVID-19. Three patients with hyperkalemia were excluded for further analysis whereas 171 patients with normokalemia were selected as control group. Hypokalemia (K <3.5 mEq/L) was detected in 119 patients (41%) and serum potassium ranged from 2.4 mEq/l to 3.5 mEq/l with a mean average of 3.1 (SD±0.18) mEq/l.

Hypokalemia was mild (3.4-3 mEq/l) in the majority of the cases (90.7%). It was associated with hypocalcemia (P=0.001) and a lower level of serum magnesium (P=0.028) compared to normokaliemic patients. Patients with hypokalemia had a longer follow-up (P=0.002) compared to normokaliemic patients likely for a severe course of COVID-19, as showed by a statistically significant higher SOFA score (P=0.014) at the admission. Mean serum creatinine of hypokalemic patients was 0.9±0.9 mg/dl. Demographics and clinical manifestations of patients are reported in Table 1 and 2.

**Table 1.**
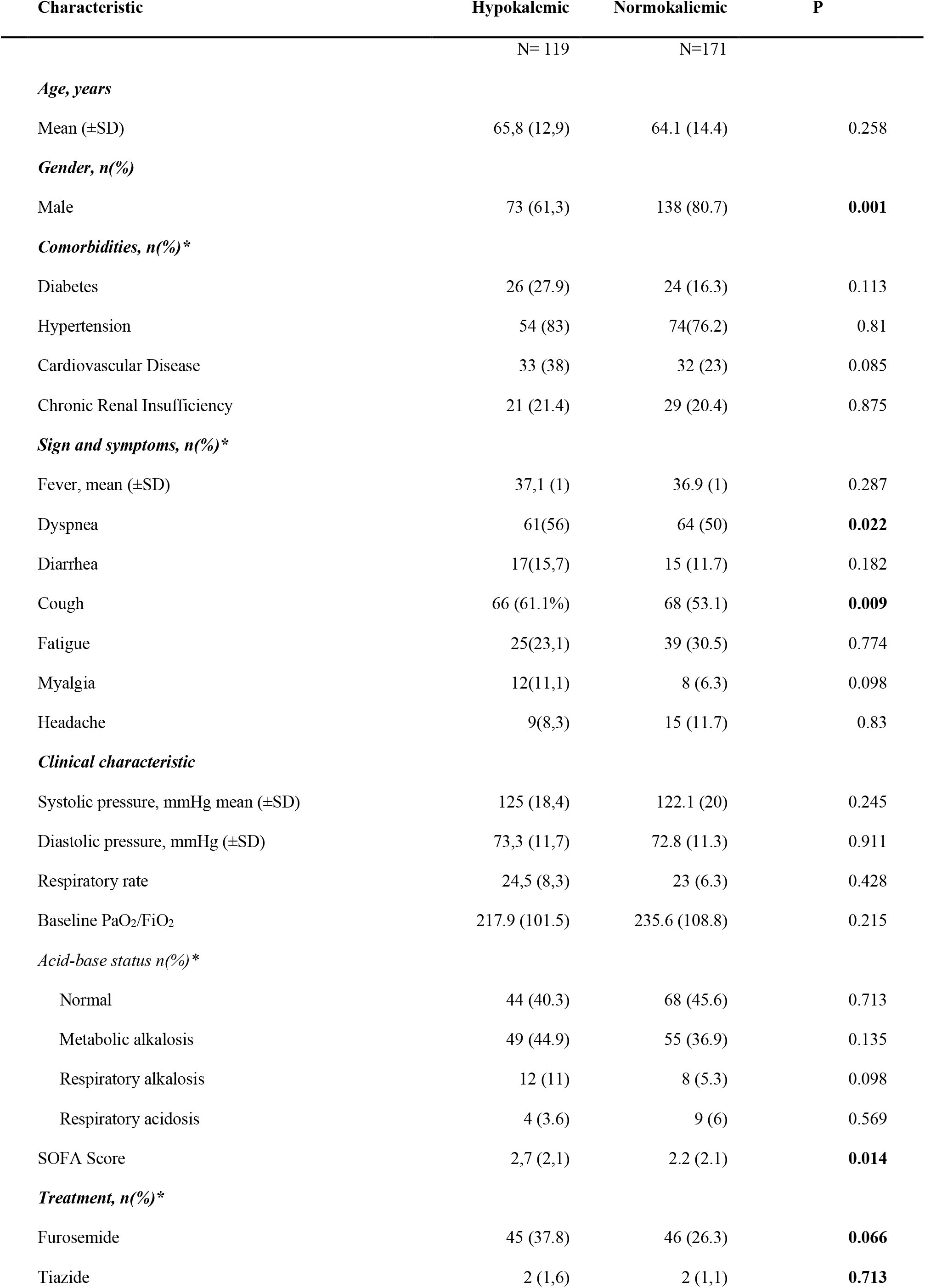
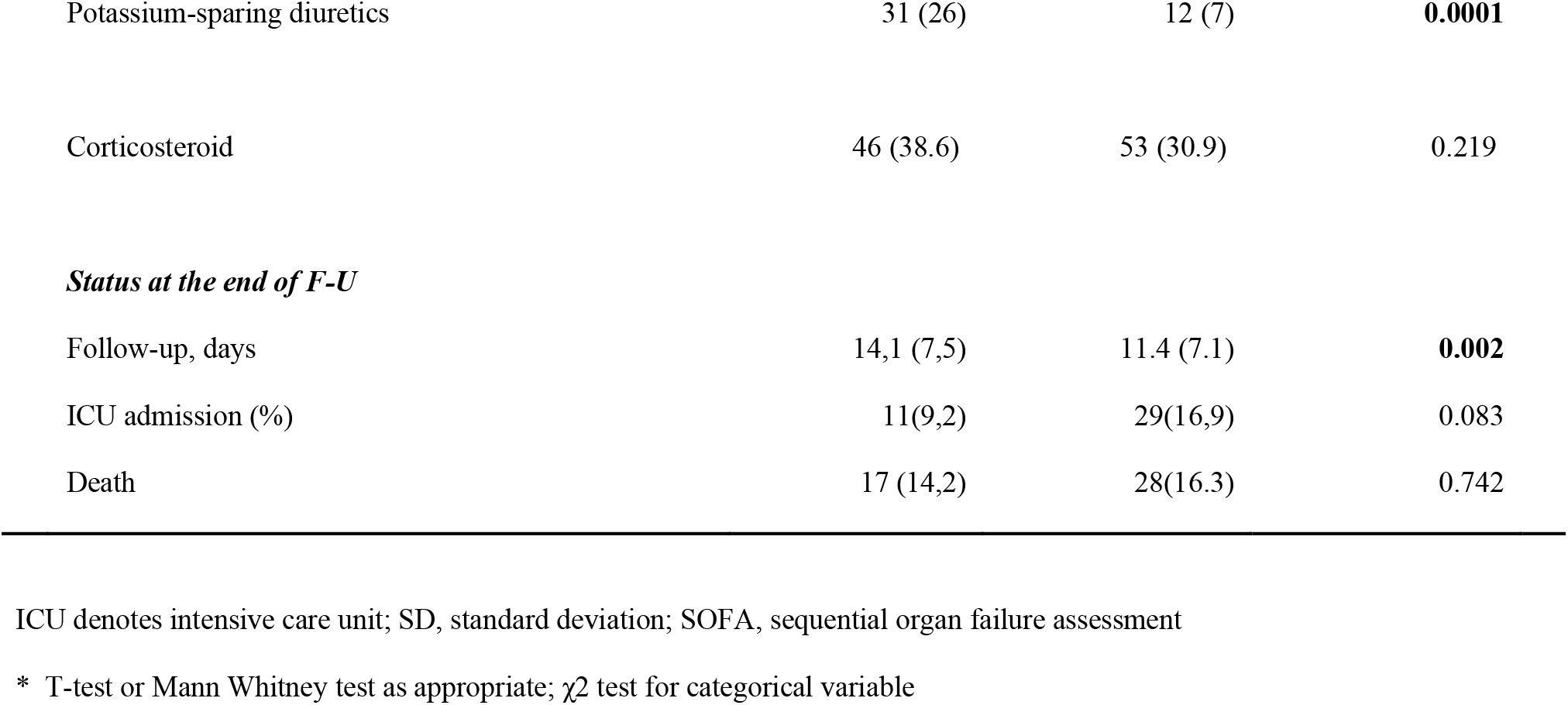
Demographic and clinical characteristics of hypokalemic (K<3.5; mEq/L) and normokaliemic (K 3.5-5.3 mEq/L) COVID-19 patients.

| Characteristic | Hypokalemic | Normokaliemic | P |
| --- | --- | --- | --- |
|  | N= 119 | N=171 |  |
| ***Age, years*** |  |  |  |
| Mean (±SD) | 65,8 (12,9) | 64.1 (14.4) | 0.258 |
| ***Gender, n(%)*** |  |  |  |
| Male | 73 (61,3) | 138 (80.7) | **0.001** |
| ***Comorbidities, n(%)\**** |  |  |  |
| Diabetes | 26 (27.9) | 24 (16.3) | 0.113 |
| Hypertension | 54 (83) | 74(76.2) | 0.81 |
| Cardiovascular Disease | 33 (38) | 32 (23) | 0.085 |
| Chronic Renal Insufficiency | 21 (21.4) | 29 (20.4) | 0.875 |
| ***Sign and symptoms, n(%)\**** |  |  |  |
| Fever, mean (±SD) | 37,1 (1) | 36.9 (1) | 0.287 |
| Dyspnea | 61(56) | 64 (50) | **0.022** |
| Diarrhea | 17(15,7) | 15 (11.7) | 0.182 |
| Cough | 66 (61.1%) | 68 (53.1) | **0.009** |
| Fatigue | 25(23,1) | 39 (30.5) | 0.774 |
| Myalgia | 12(11,1) | 8 (6.3) | 0.098 |
| Headache | 9(8,3) | 15 (11.7) | 0.83 |
| ***Clinical characteristic*** |  |  |  |
| Systolic pressure, mmHg mean (±SD) | 125 (18,4) | 122.1 (20) | 0.245 |
| Diastolic pressure, mmHg (±SD) | 73,3 (11,7) | 72.8 (11.3) | 0.911 |
| Respiratory rate | 24,5 (8,3) | 23 (6.3) | 0.428 |
| Baseline PaO<sub>2</sub>/FiO<sub>2</sub> | 217.9 (101.5) | 235.6 (108.8) | 0.215 |
| ***Acid-base status n(%)\**** |  |  |  |
| Normal | 44 (40.3) | 68 (45.6) | 0.713 |
| Metabolic alkalosis | 49 (44.9) | 55 (36.9) | 0.135 |
| Respiratory alkalosis | 12 (11) | 8 (5.3) | 0.098 |
| Respiratory acidosis | 4 (3.6) | 9 (6) | 0.569 |
| SOFA Score | 2,7 (2,1) | 2.2 (2.1) | **0.014** |
| ***Treatment, n(%)\**** |  |  |  |
| Furosemide | 45 (37.8) | 46 (26.3) | **0.066** |
| Tiazide | 2 (1,6) | 2 (1,1) | **0.713** |
| Potassium-sparing diuretics | 31 (26) | 12 (7) | **0.0001** |
| Corticosteroid | 46 (38.6) | 53 (30.9) | 0.219 |
| ***Status at the end of F-U*** |  |  |  |
| Follow-up, days | 14,1 (7,5) | 11.4 (7.1) | **0.002** |
| ICU admission (%) | 11(9,2) | 29(16,9) | 0.083 |
| Death | 17 (14,2) | 28(16.3) | 0.742 |

ICU denotes intensive care unit; SD, standard deviation; SOFA, sequential organ failure assessment
\* T-test or Mann Whitney test as appropriate; $\chi^2$ test for categorical variable

**Table 2.** Main lab examinations of hypokalemic and normokaliemic patients

| *Lab test, Mean (±SD)* | **Hypokalemic** | **Normokalemic** | ***P value\**** |
| --- | --- | --- | --- |
| Hemoglobin, g/l | 12,1 (1,6) | 12.8 (1.793) | **0.001** |
| White cells, mm3 | 7393 (5044) | 8156.6 (8228.1) | 0.750 |
| Platelets, 10<sup>9</sup>/l | 252,3 (119,3) | 243.2 (116.1) | 0.482 |
| Glycemia | 105,2 (35,9) | 110.6 (54.4) | 0.656 |
| Potassium, mmol/l | 3,1 (0,1) | 4.0 (0.3) | **< 0.0001** |
| Sodium, mmol/l | 137.4 (3,9) | 137.2 (3.6) | 0.818 |
| Calcium, mg/l | 8,3 (0,5) | 8.7 (0.6) | **0.001** |
| Magnesium | 2,1 (0,3) | 2.6 (0.3) | **0.028** |
| Chloride, mmol/l | 99.6 (4,5) | 100.2 (4.4) | 0.509 |
| Creatinine, mg/l | 0,9 (0,9) | 1.1 (1.3) | **< 0.0001** |
| Urea, mg/dl | 55,31 (47,8) | 52 (30.3) | 0.564 |
| Albumin, gr/dl | 3,2 (0,5) | 3.1 (05) | 0.519 |
| Urine K<sup>+</sup> (spot) | 30,1 (16,7) | 38.1 (21.8) | 0.229 |
| Urine Creatinine (spot) | 144,4 (72,4) | 110.1 (62.1) | 0.079 |
| Urine protein-to-creatinine | 0,5 (0,5) | 0.4(0.31) | 0.300 |
| D-dimer, mg/l | 2478 (4070.8) | 3045.7 (5718.3) | 0.826 |
| Alanine amino-transferase, U/l | 45,9(37,2) | 109.65 (761.7) | 0.371 |
| Lactate dehydrogenase, U/l | 687,2 (310) | 812.2 (1914.1) | 0.348 |
| Ferritin, mg/dl | 867 (416.7) | 856.4 (824.1) | 0.373 |
| Bilirubin, mg/l | 0.7 (0.5, 0.3) | 0.7 (0.7) | 0.301 |
| C-reactive protein, mg/l | 10,2 (9,9) | 9.8 (7.6) | 0.524 |

\*T-test or Mann Whitney test as appropriate

**Table 3A.**
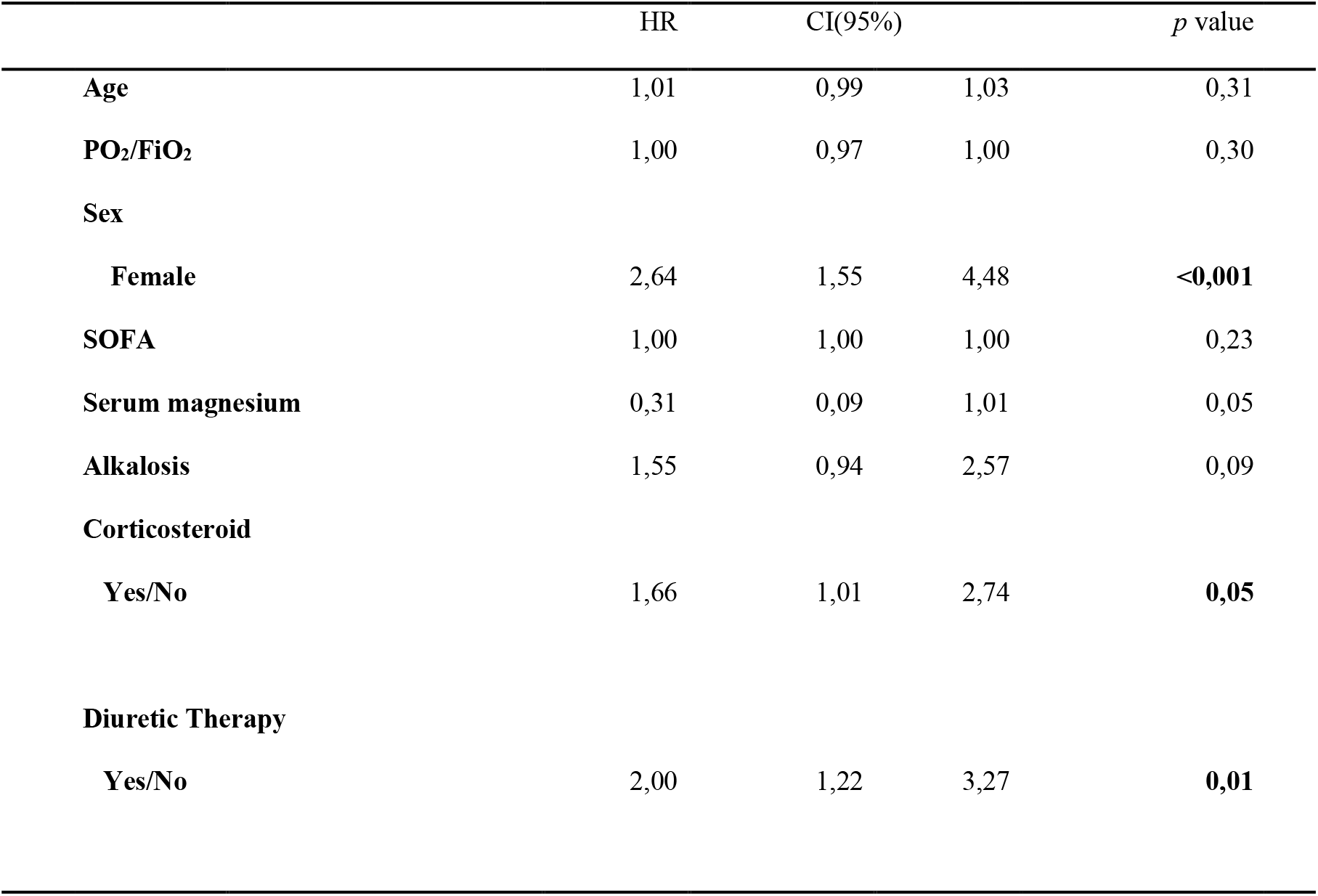
Predictive factors of hypokalemia revealed by univariate logistic regression analysis

**Table 3B.** Multivariate regression model of predictive factors of hypokalemia

|  | HR | CI(95%) |  | *p* value |
| --- | --- | --- | --- | --- |
| **Alkalosis** | 1,50 | 0,89 | 2,53 | 0,128 |
| **Diuretic Therapy** | 1,94 | 1,08 | 3,48 | **0,027** |
| **Sex** |  |  |  |  |
| **Female** | 2,44 | 1,36 | 4,37 | **0,003** |
| **Corticosteroid** | 1,25 | 0,69 | 2,27 | 0,467 |

### Potassium excretion

Urine K-to-creatinine ratio was measured only in a minority of hypokalemic patients (n=45;36.1%). Urine K-to-creatinine ratio >1.5 mEq/mmol, expression of high urinary excretion of a potassium[13], was documented in 43 (95.5%) patients. Twenty-three patients (53.4%) with high urinary potassium excretion were on diuretic therapy (furosemide) and 10 (23.3%) were on corticosteroids at the time of serum K measurement. The remaining 10 patients (23.3%) with augmented kaliuresis had normal serum level of magnesium and 90% had low sodium excretion (FENa< 1%) (Fig.1). Acid-base status of these ten patients showed that 70% had metabolic alkalosis, 10% respiratory alkalosis, 10% respiratory acidosis and the remaining 10% a normal acid-base status

**Figure1.**
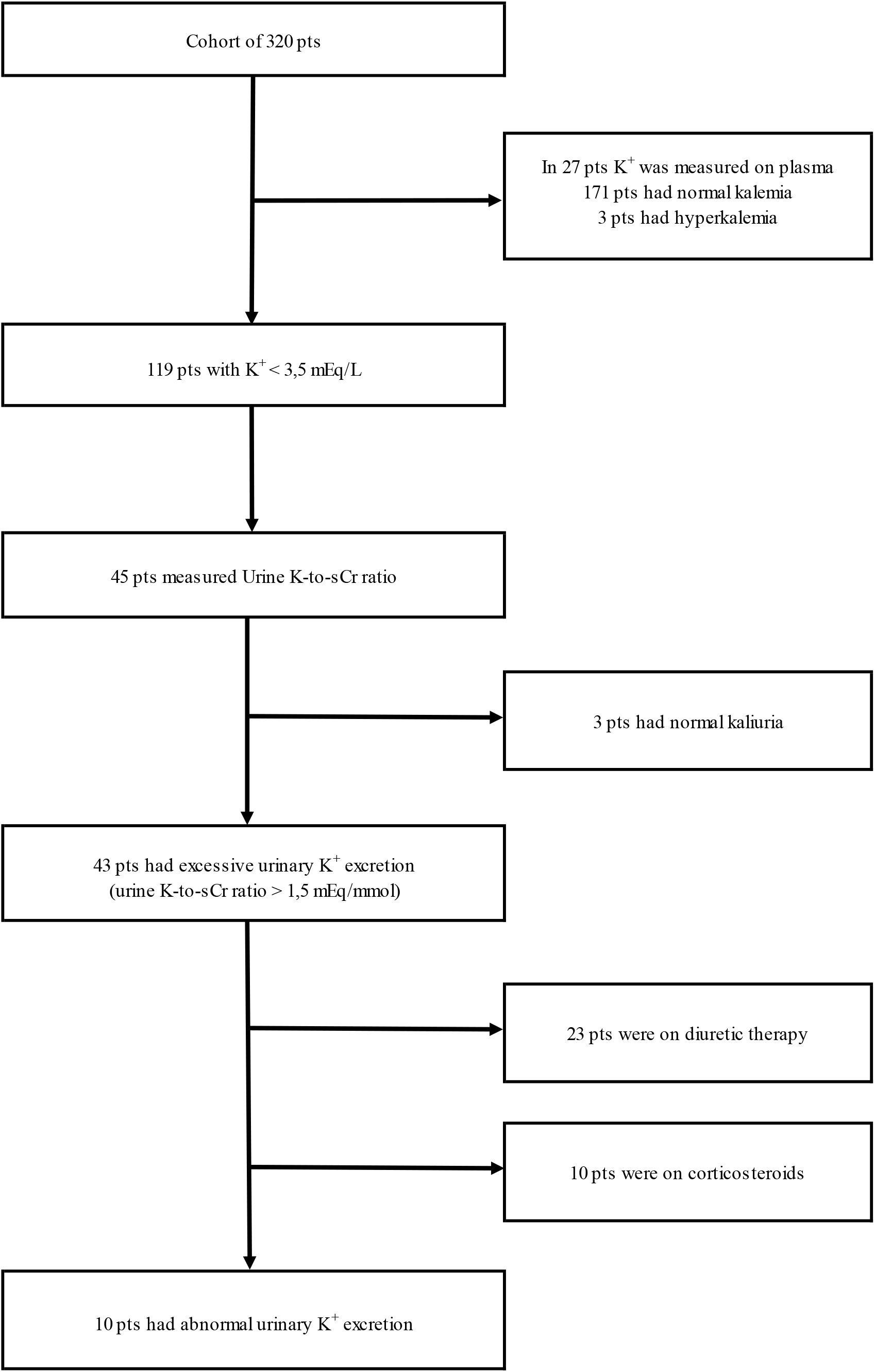
Flow chart of patients’ selection.

### Therapy in hypokalemic patients

Potassium supplements were administrated to correct hypokalemia (Table 2). Salts of potassium (KCl, 600mg) were administered via the oral route in 28 (23.5%) patients and IV in 7 (5.8%) patients, both oral route and iv were used in one (0.8%) patient.

Intravenous magnesium sulfate was administered in 22 (18.4%) patients for hypomagnesemia Potassium-sparing diuretics were largely administered among hypokalemic patients (26 %) compared to normokalemia (7%), likely for its pharmacological action targeted to maintain a normal kalemia and prevent hypokalemia (Table 2). Forty-five (37.5%) patients were on diuretic therapy with furosemide and two (1.6%) with thiazide at the time of diagnosis with hypokalemia. Hypertension (57.4%), cardiovascular disease (38.2%) and renal impairment (31.9%) were the potential causes of diuretic use in these patients.

### Risk factor and Outcome

Univariate analysis showed that female gender, corticosteroids and diuretics were significantly associated with hypokalemia in our cohort of patients. Multivariate regression model analysis showed that diuretic therapy (P=0.027; HR 1.94; 95% CI 1.08-3.48) and female sex (P=0.003; HR 2.44; 95% CI 1.36-4.37) were significant risk factors for hypokalemia. Survival regression analysis showed that hypokalemia adjusted for sex, age and SOFA score was not associated with intensive care admission (P=0.131, 95% CI 0.228-1.212) and in-hospital mortality (P=0.474; 95% CI 0.170-1.324).

## Discussion

The results of our study showed that hypokalemia is a common electrolytic abnormality among patients with COVID-19. Hypokalemia has been found in 41% of patients during hospitalization for severe symptoms of SARS-CoV-2 infection. The prevalence and causes of hypokalemia are unknown until now, as this phenomenon has been only mentioned as a potential manifestation of COVID-19[7]. However, there is a general concern about the risk of hypokalemia to trigger life-threatening arrhythmia in patients affected by SARS-CoV-2[7,8].

Diuretic therapy along with female sex was significantly associated with hypokalemia in our study-cohort. Diuretics are pharmacological agents largely used in the general population to treat hypertension and prevent fluid overload. The slightly higher prevalence of hypertension, cardiovascular disease and renal impairment in hypokaliemic than normokaliemic patients may be a plausible explanation of the higher use of diuretics in this group of patients. Diuretics are widely known to cause hypokalemia and other electrolyte imbalances with long-term administration [14], as they act by inhibiting reabsorption of electrolytes in the renal tubules. This side effect was likely the major determinant of other electrolyte disturbances in our patients. Hypokalemia was indeed associated with hypocalcemia and a lower level of serum magnesium compared to normokaliemic patients. Although the female sex appears poorly interlinked with hypokalemia, the first experimental studies conducted in the 50s [15] and then confirmed in the 90s [11] showed that women, especially aged ones, have less exchangeable body potassium than other subset of the popualtion. The women are, therefore, at high risk to develop hypokalemia because they have depleted deposits of potassium due to their different body composition, characterized by less amount of extracellular water compared to men.

Hypokalemia is a frequent electrolyte disturbance in hospitalized patients. Multiple mechanisms underlying this condition include diuretic therapy, gastrointestinal loss, anorexia or alkalosis[22]. More rarely, congenital or acquired tubular defect, side effects of drugs or tubular ischemia induce tubular potassium losses due to the disruption of renal electrolytes handling[23,24]. Hyperaldosteronism, primitive or as a consequence of activation of the renin-angiotensin system, is also known to stimulate urinary potassium excretion. In the setting of COVID-19 infection, several adjunctive conditions can disrupt potassium homeostasis during hospitalization. Respiratory alkalosis due to hypoxia-driven hyperventilation may provoke transcellular shifts with increased intracellular uptake, and anorexia, as a consequence of continuative use of face mask or ventilation helmet or status of severe illness, may lead to a decrease in potassium intake; diarrhea due to medication (e.g., lopinavir/ritonavir) or the cytopathic effect of the virus on the gastrointestinal cells may be a cause of potassium losses.

Moderate and severe hypokalemia has widely been associated with potentially life-threatening complications such as cardiac dysrhythmias, paralysis and rhabdomyolysis [16]. Underlying cardiovascular disease [17,18] and chronic kidney disease [19,20] are widely recognized as a poor prognostic factor for mortality among hypokalmic patients.

The findings of our study show that patients with hypokalemia had a statistically longer hospitalization than controls. Likely the higher rate of respiratory symptoms (dyspnea and cough) and the worse SOFA score (2.7 vs 2.2) at admission reflected a more severe systemic inflammatory response leading to a prolonged hospital stay than normokaliemic patients. Nevertheless, we did not notice that hypokalemia was not a poor prognostic factor for intensive care admission (P=0.131, 95% CI 0.228-1.212) and hospital mortality (P=0.474; 95% CI 0,170-1,324). Likely the mild severity of hypokalemia (90.7% had serum levels of potassium between 3.4 and 3 mEq/L) may explain the null effect of hypokalemia on major outcomes in our cohort of patients. Despite these reassuring data, serum potassium should be carefully monitored in this vulnerable subset of the population for the risk of potentially lethal arrhythmia. Hypokalemia may further prolong QT interval in COVID-19 patients with abnormal QT interval due to the large use of off-label agents for SARS-CoV-2 infection such as azithromycin and hydroxychloroquine.

To date, few reports document an interplay between hypokalemia and coronavirus infection. Hypokalemia has been reported during the SARS-CoV-1 outbreak in Canada. Booth et al**[9]** described that 62 out of 144 (43%) patients had mild hypokalemia (mean, 3.2 mEq/l) during hospitalization. Hypokalemia was not isolated and occurred with other electrolyte abnormalities such as hypomagnesemia and hypophosphatemia. The authors were unable to establish clear evidence for electrolyte imbalance and suggested that it may be a clinical manifestation of the infection or a nephrotoxic effect of ribavirin or other medicaments on renal electrolytes handling. The abstract of a Chinese article, reported hypokalemia as a side effect of glucocorticoid administration in SARS-CoV-1 infected patients**[10]**. These agents through activation of the mineralocorticoid receptor on the renal tubular cells could have increased urinary potassium excretion**[25]**.

In an attempt to clarify tubular potassium handling in COVID-19 we investigated the magnitude of potassium secretion in a minority of hypokalemic patients. Urine K-to-creatinine ratio was augmented in 95.5% of patients who underwent urinary examination (n=45), expression of pathological tubular potassium handling. After the exclusion of about three-fourths of urinary samples because of intercurrent diuretic and corticosteroid therapy, we found that in the small proportion of remaining patients (n=10, 23.3%), urinary examination was characterized by an augmented urinary potassium excretion in the absence of concurrent causes (normal serum magnesemia). Interestingly, most of them (70%) had metabolic alkalosis on arterial blood gas analysis. Based on these data, the etiology of hypokalemia of COVID patients seems multifactorial and a side effect of diuretic and corticosteroid therapy. However, we cannot overlook the inappropriate loss of urinary potassium occurring in a small number of patients in the absence of apparent pharmacological interferences. Two hypotheses should be enunciated regarding this issue. First, SARS-COV-2 may mediate a direct viral cytopathic effect on kidney tubular proximal cells with loss of renal tubular cell integrity and function. In support of this hypothesis, the virus has been identified in urine samples in patients with severe COVID-19[26], as ACE 2, virus entry receptor, is largely expressed within the kidney, especially in tubular cells[27]. The virus may cause diffuse acute proximal tubular injury characterized by loss of apical brush border and vacuolation[11]. The second hypothesis relies on hyperaldosteronism due to the abnormal activation of the renin-angiotensin system as a result of ACE2 disruption following cell infection[28]. The findings of metabolic alkalosis, low sodium execration corroborate this theory, but the lack of data on serum renin and aldosterone levels cannot corroborate our hypothesis.

Several other limitations of this study must be enunciated. First, we were unable to trace hypokalemia-related clinical manifestations despite the completeness of the electronic chart. Second, the restricted group of patients who underwent urinary examination has reduced the accuracy of analysis to identify the causes of hypokalemia. However, despite the small sample, we have collected comprehensive data on magnesium level, urine sodium and acid-base status.

This study is the first report describing in detail the clinical characteristic of hypokalemic patients affected by SARS-CoV-2. Although urine examination was available only in a small number of patients, we traced the potential causes of hyperkalemia, including diuretic therapy, acid-base disorder and corticosteroid therapy, in a cohort of hospitalized patients. Our results support the need to frequently measure the level of serum potassium along with urine K-to-creatinine ratio or 24h potassium excretion in COVID-19 patients in order to improve the management of these patients. Based on the magnitude of the urinary loss, potassium should be supplemented per os or IV, and a careful assessment of the ECG should be performed, especially in concomitance of potentially arrhythmogenic drugs. Further studies are required to investigate the hypothesized cytopathic effect of SARS-CoV-2 on renal tubular cells and other secondary injury mechanisms such as hypoxia and cytokine storm.

## Conclusions

Hypokalemia was a frequent electrolyte disorder in our hospitalized patients with COVID-19. It was highly associated with urinary loss due principally to iatrogenic causes. In a small proportion of patients (22.2%), the etiology of tubular potassium loss remained elusive. Generally, hypokalemia was mild, not associated with poor outcomes and was corrected with oral supplementation of potassium. Although hypokalemia was not associated with mortality in our cohort of patients, it may be a life-threatening condition (e.g., long QT) if it remains untreated.

## Data Availability

Data are available upon reasonable request

